# Nepalese version of Douleur Neuropathique 4 (DN4) questionnaire for Assessment of Neuropathic pain: A Validation Study

**DOI:** 10.1101/2021.09.17.21263734

**Authors:** Bigen M Shakya, Anil Shrestha, Amod K Poudyal, Ninadini Shrestha, Binita Acharya, Renu Gurung, Sujata Shakya

## Abstract

**Objective:** This study aimed to translate and validate DN4 questionnaire into Nepalese version.

**Design:** An observational study design was adopted.

**Setting:** A tertiary level teaching hospital of Kathmandu, Nepal

**Participants:** We included 166 purposively selected patients visiting pain clinics of the hospital over one year time

**Methods:** The Nepalese version of the DN4 questionnaire was used to detect neuropathic pain among the chronic pain patients of the hospital. The English version of the questionnaire was translated into Nepali based on the standard guideline with the help of linguistic experts. The patients diagnosed with nociceptive or neuropathic pain were interviewed twice in two weeks interval. We analyzed test-retest reliability and strength of the test by using Intra-class correlation coefficient (ICC) and Receiver Operating Characteristics (ROC) Curve, respectively. Internal Consistency reliability was assessed using Cronbach’s alpha (∞). Diagnostic accuracy was assessed through measures like sensitivity, specificity, positive and negative predictive values, and positive and negative likelihood ratio.

**Results:** The study showed a good test-retest reliability (ICC=0.877) and internal consistency reliability (∞=0.710). The AUC were 0.932 (0.894-0.971) for the first test, and 0.955 (0.921-0.990) for the second test. The sensitivity and specificity values were found highest at the 4 cut-off point (4 score out of 10), that are 75% and 95.3% for test 1, and 76.2% and 98.8% for test 2. Similarly positive and negative predictive values are 93.8% and 80.4% respectively for the first test and 98.4% and 81.7% respectively for the second test.

**Conclusions:** The Nepalese version of DN4 questionnaire is a valid and reliable tool for the diagnosis of neuropathic pain. This can be used for screening neuropathic and non-neuropathic pain in clinical as well as epidemiological settings.

**Article Summary:** *Strengths and Limitations of this Study:*

- This study validated Nepalese version of DN4 questionnaire, so, it can be used as a standard tool to assess neuropathic pain among the Nepalese population.
- As interview was conducted with the patients, this might minimize the reliability and validity issue.
- This questionnaire is only applicable to those who can communicate properly in Nepalese language.
- There can be problem of understanding among the participants as few words do not have exact Nepali words with the same meaning.

## Introduction

Neuropathic pain is defined as pain caused by a lesion or disease of the somatosensory nervous system (1). The diagnosis of neuropathic pain is complex, and it is based on clinical history, physical examination, and other advanced investigations. Since neuropathic pain is one of the prevalent health conditions, several questionnaires have been developed to facilitate its detection. They include Neuropathic pain Scale (2), Pain detect (3), Leeds Assessment of Neuropathic Symptoms and Signs (LANSS) (4), Douleur Neuropathique 4 (DN4). These screening tools consist of structured questions and may include a simple clinical examination. The detection of neuropathic pain can be done with a high degree of sensitivity and specificity with the aforementioned tools (5). Moreover, these tools can be used by a specialist as well as any health person (6).

Due to its popularity, the DN4 questionnaire has been translated and validated into various languages such as Hindi (7), Thai (8), Japanese (9), Korean (10), Spanish (11), Greek (12), Portuguese (13), Turkish (14) and so on. It was originally developed by French Neuropathic pain group in 2005 (15). This screening tool is simple to use and can be introduced in daily practice as well for epidemiological studies.

Translation of the standard questionnaire should preserve the meaning and intent of the original item, and its validity and reliability must be maintained. The process of translation and its adaptation to different cultural groups are demanding and time consuming. Without validation, translation of a questionnaire may lead to misleading conclusions, either in clinical practice and/or epidemiological studies. There exist guidelines for the translation and validation of tools (16). Some of the standard tools, like Numerical Pain Rating Scale, have already been translated and validated in Nepali language (17). Our aim is to translate and validate the DN4 questionnaire according to the standard guideline. Once validated, this tool can be used as a screening tool by any health personnel for the diagnosis of neuropathic pain.

## Methods

### Study Design

We conducted a prospective observational study among patients with chronic pain attending the pain clinic of a tertiary level teaching hospital, Kathmandu, Nepal, over a period of one year from February 2019 to January 2020.

### Participants Selection

For the selection of participants, we adopted nonprobability purposive sampling technique and included adult patients (18 years and above) who attended the pain clinic with chronic pain (pain of duration more than 3 months) or who were referred from other departments for further evaluation of chronic pain. Only those patients who could communicate in Nepalese language were selected for the study. The excluded patients were those who had fibromyalgia, phantom pain, headache, chronic visceral pain, cancer pain, and severe depression.

Due to the lack of consensus regarding the calculation of the sample size in validation studies, the suggested ratio of subject to item ranges from 2-20 (18). The total sample size for this study was 166, which was taken to make the subject-to-item ratio 16:1.

The ethical approval for the research was obtained from the Institutional Review Committee of Institute of Medicine, Tribhuvan University [338(6-11)E^2^/075/76]. Written informed consent was taken from the participants before data collection.

### Data Collection

We used Nepalese version of the DN4 questionnaire among the chronic pain patients by using interview technique to detect the neuropathic pain.

#### DN4 questionnaire

The DN4 questionnaire consists of 10 items, in which 7 items are related to pain characteristics and 3 items are related to findings on physical examination of the painful areas. The cut-off value for the diagnosis of neuropathic pain is 4/10. The area under the curve for the total score of French version of the questionnaire is 0.92. A cut-off score of 4 resulted in the highest percent of sensitivity (82.9%) and specificity (89.9%). The inter-rater reliability (Cohen Kappa coefficient) is between 0.70 and 0.96. (15)

We adopted guidelines given by Sperber AD for translation process.(16)

#### Phase 1: Translation and back-translation

The English version of DN4 questionnaire was translated into Nepalese version independently by three individuals, two linguistic experts, and one neurologist who had experience in management of chronic pain. These three individuals along with the principal investigator then discussed issues during translation process and formed a preliminary draft of Nepalese version of the questionnaire. The preliminary draft so formed was back translated to English version by another translation expert who was not aware of the previous translation. The committee of three pain specialists (Anaesthesiologists trained in management of chronic pain), along with the forward and backward translators and a research expert discussed and reviewed the items of all versions of translation and compared with the original English version for semantic, experiential and conceptual equivalence. Finally, the pre-final Nepalese version of DN4 was formed.

#### Phase 2: Pretesting and modification

Pretesting of the pre-final Nepalese version was done among 25 patients with neuropathic pain attending a private pain clinic situated in Kathmandu, Nepal. During the interview, the patients were specifically asked “Do you understand about items listed in the questionnaire?”. Depending on the results of the pretest, the Nepali word for ‘cold pain’ was rephrased, and the final draft of the Nepalese version of Dn4 questionnaire was prepared.

Face validity of the questionnaire was maintained by pretesting the pre-final version of the translated Nepalese DN4 questionnaire among a small group of respondents. We found that all of the pain descriptors were well understood by the respondents excluding the word ‘cold pain. Various possible words were explored which is closest to the word ‘cold pain’. Finally, the word “□□□□ □□□□ □□□□□ □□□□ □□□□ □□□□□□□ □□□□□” was selected as the most appropriate word. The new translated word was retested in a small group of respondents, which showed the satisfactory results.

Content validity of the questionnaire was established by forming a panel of experts containing a methodologist, a statistician and two pain specialists. The translated Nepalese version of the questionnaire was compared with the previous English version and discussed among the experts. The similarity of interpretability of words, phrases, and sentences were analyzed, and the questionnaire was examined to find out whether the items adequately measure the intended constructs and are sufficient to measure different domains. The translation was revised based on consensus of all.

### Test Methods

We divided the patients with chronic pain into two groups (neuropathic and nociceptive) after examination by a pain specialist in the pain clinic. The diagnosis of neuropathic pain was made through the patient’s history, clinical examination, and/or confirmatory tests (if applicable) based on the guidelines provided by International Association for the Study of Pain (IASP) which is the internationally accepted guideline (19). A pain specialist, who was not involved in the segregation of patients, then performed interviews on the same day using the final corrected Nepali version of DN4 questionnaire. Those patients were interviewed again within two weeks using the same questionnaire during the follow-up. We considered the cutoff value for leveling neuropathic pain by DN4 questionnaire as 4/10 and above. This cutoff was also originally used in French version (15).

### Statistical Analysis

We entered the data into Microsoft Excel 2013 and then exported to SPSS version 16 for statistical analysis. Under descriptive statistics, mean, frequency and percentage were calculated. To compare the score of DN4 questionnaire with the nature of pain, chi-square test was performed. To assess the internal consistency reliability, Cronbach’s alpha was calculated. Cronbach’s alpha of ≥ 0.7 is considered adequate. (20) We used test-retest reliability to assess the correlation between two episodes of the test and derived Receiver Operating Characteristic (ROC) Curve for analyzing the discriminative ability of the test. Similarly, different diagnostic values to detect neuropathic pain like sensitivity, specificity, positive and negative predictive value, and positive and negative likelihood ratio were calculated. Any missing or indeterminate test results were excluded from the study.

### Patient and Public Statement

No patient or public involvement was done during design, conduct, reporting or dissemination of the research. The patients attending pain clinic were involved as participants after obtaining written informed consent.

## Results

The total participants attending the pain clinic during the period of the study was 216. Out of these, 50 patients were excluded due to various reasons like cancer, headache, pain less than 3 months, phantom pain, and visceral pain. Thus, the total number of eligible participants enrolled in this study was 166.

The general characteristics of the respondents are presented in Table 1. The mean ages of the patients with nociceptive pain and neuropathic pain were 49.03±15.1 and 52.79±14.8 years respectively. More than two-third (67.4%) were females.

**Table 1.**
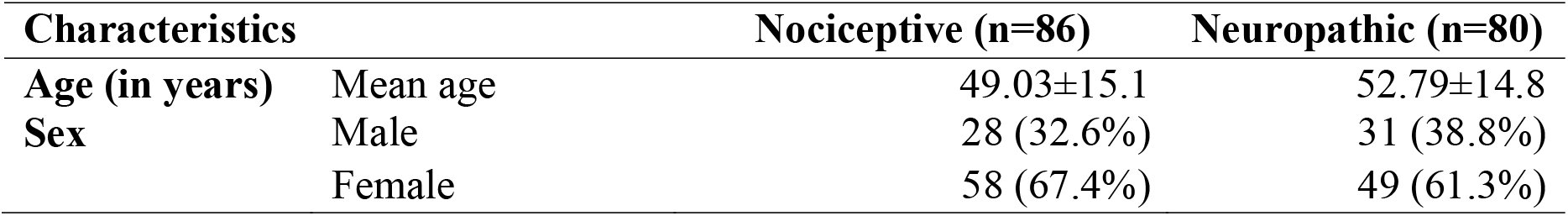
Baseline Characteristics.

The most common symptoms in patients with neuropathic pain were tingling sensation (75.0%), burning pain (72.5%), needle sensation (71.3%), and electrical pain (71.3%) (Table 2). These symptoms were found significantly more among patients with neuropathic pain compared to those with nociceptive pain (p<0.001).

**Table 2:** Comparison of Items of DN4 Questionnaire between Neuropathic and Nociceptive conditions.

| Items | Nociceptive (n=86) |  |  |  | Neuropathic (n=80) |  |  |  | p value* |
| --- | --- | --- | --- | --- | --- | --- | --- | --- | --- |
|  | Absent |  | Present |  | Absent |  | Present |  |  |
|  | n | % | n | % | n | % | n | % |  |
| **Burning** | 61 | 70.9 | 25 | 29.1 | 22 | 27.5 | 58 | 72.5 | <0.001 |
| **Cold pain** | 79 | 91.8 | 7 | 8.1 | 66 | 82.5 | 14 | 17.5 | 0.7 |
| **Electric pain** | 74 | 86.0 | 12 | 14.0 | 23 | 28.8 | 57 | 71.3 | <0.001 |
| **Tingling** | 66 | 76.7 | 20 | 23.3 | 20 | 25.0 | 60 | 75.0 | <0.001 |
| **Needle sensation** | 56 | 65.1 | 30 | 34.9 | 23 | 28.8 | 57 | 71.3 | <0.001 |
| **Numbness** | 81 | 94.2 | 5 | 5.8 | 45 | 56.3 | 35 | 43.8 | <0.001 |
| **Itching** | 85 | 98.8 | 1 | 1.2 | 66 | 82.5 | 14 | 17.5 | <0.001 |
| **Decreased sense on touch** | 84 | 97.7 | 2 | 2.3 | 53 | 66.3 | 27 | 33.8 | <0.001 |
| **Decreased sense on pin pick** | 86 | 100.0 | 0 | 0 | 56 | 70.0 | 24 | 30.0 | <0.001 |
| **Increased pain on brushing** | 83 | 96.5 | 3 | 3.5 | 55 | 68.8 | 25 | 31.3 | <0.001 |

\*Chi-square test

Table 3 compares the pain assessment using DN4 questionnaire with the diagnosis of pain. It shows a highly significant association between the two scores of DN4 questionnaire and the actual diagnosis of pain (p<0.001).

**Table 3.** Comparison of the score of DN4 questionnaire with diagnosis of pain.

|  |  | Diagnosis of pain |  |  |  | p value* |
| --- | --- | --- | --- | --- | --- | --- |
|  |  | Nociceptive (n=86) |  | Neuropathic (n=80) |  |  |
|  |  | n | % | n | % |  |
| DN4 first time | Negative | 82 | 80.4 | 20 | 19.6 | <0.001 |
|  | Positive | 4 | 6.3 | 60 | 93.8 |  |
| DN4 second time | Negative | 85 | 81.7 | 19 | 18.3 | <0.001 |
|  | Positive | 1 | 1.6 | 61 | 98.4 |  |

\*Chi-square test

### Internal Consistency Reliability (Chronbach’s alpha)

The Cronbach’s alpha (∞) coefficient of the entire DN4 questionnaire is found to be 0.710, indicating adequate internal consistency. The coefficient value was not much difference by removing any of the items in the questionnaire. Thus, all questions were included in the DN4 questionnaire (Table 4).

**Table 4:** Internal Consistency Reliability.

| Item | Cronbach's $\alpha$ | ICC (95% CI) | Cronbach's $\alpha$<br>(if single item deleted) |
| --- | --- | --- | --- |
| Burning | 0.710 | 0.640-0.772 | 0.689 |
| Cold pain |  |  | 0.728 |
| Electric pain |  |  | 0.677 |
| Tingling |  |  | 0.656 |
| Needle sensation |  |  | 0.710 |
| Numbness |  |  | 0.664 |
| Itching |  |  | 0.701 |
| Decreased sense on touch |  |  | 0.660 |
| Decreased sense on pin pick |  |  | 0.661 |
| Increased pain on brushing |  |  | 0.719 |

ICC = Intra-class Correlation Coefficient

### Test–retest Reliability

The patients were interviewed twice using the Nepalese version of DN4 questionnaire, once during their first visit at the pain clinic, and again within 2 weeks. The test-retest reliability was measured by calculating intra-class correlation coefficient, which was found to be more than 0.8, indicating a good test-retest reliability value (Table 5).

**Table 5:** Test-retest reliability of the Nepali version of the DN4 questionnaire.

Test-retest Reliability
| | Test (Mean±SD) | Re-test (Mean±SD) | ICC (95% CI) |
| --- | --- | --- | --- |
| Retest sample (n=166) | 3.06±2.293 | 2.74±2.235 | 0.948 (0.921-0.964)* |
| NP (n=80) | 4.79±1.874 | 4.55±1.764 | 0.877 (0.807-0.921)* |
| NNP (86) | 1.45±1.233 | 1.06±0.925 | 0.858 (0.689-0.925)* |

NP = Neuropathic Pain, NNP = Non-neuropathic Pain, ICC = Intra-class Correlation Coefficient, \*p<0.001 (p value for intra-class correlation coefficient)

### Receiver Operating Characteristics Curve (ROC) Analysis

The clinical diagnosis was used as the reference for the calculation of Area under the Curve (AUC). Figure 1 represents the ROC curve for two episodes of the test using Nepali version of DN4 questionnaire. The AUCs for the two episodes of test were 0.932 with 95% CI (0.894 to 0.971) and 0.955 with 95% CI (0.921-0.990), which signifies a strong test.

**Figure 1.**
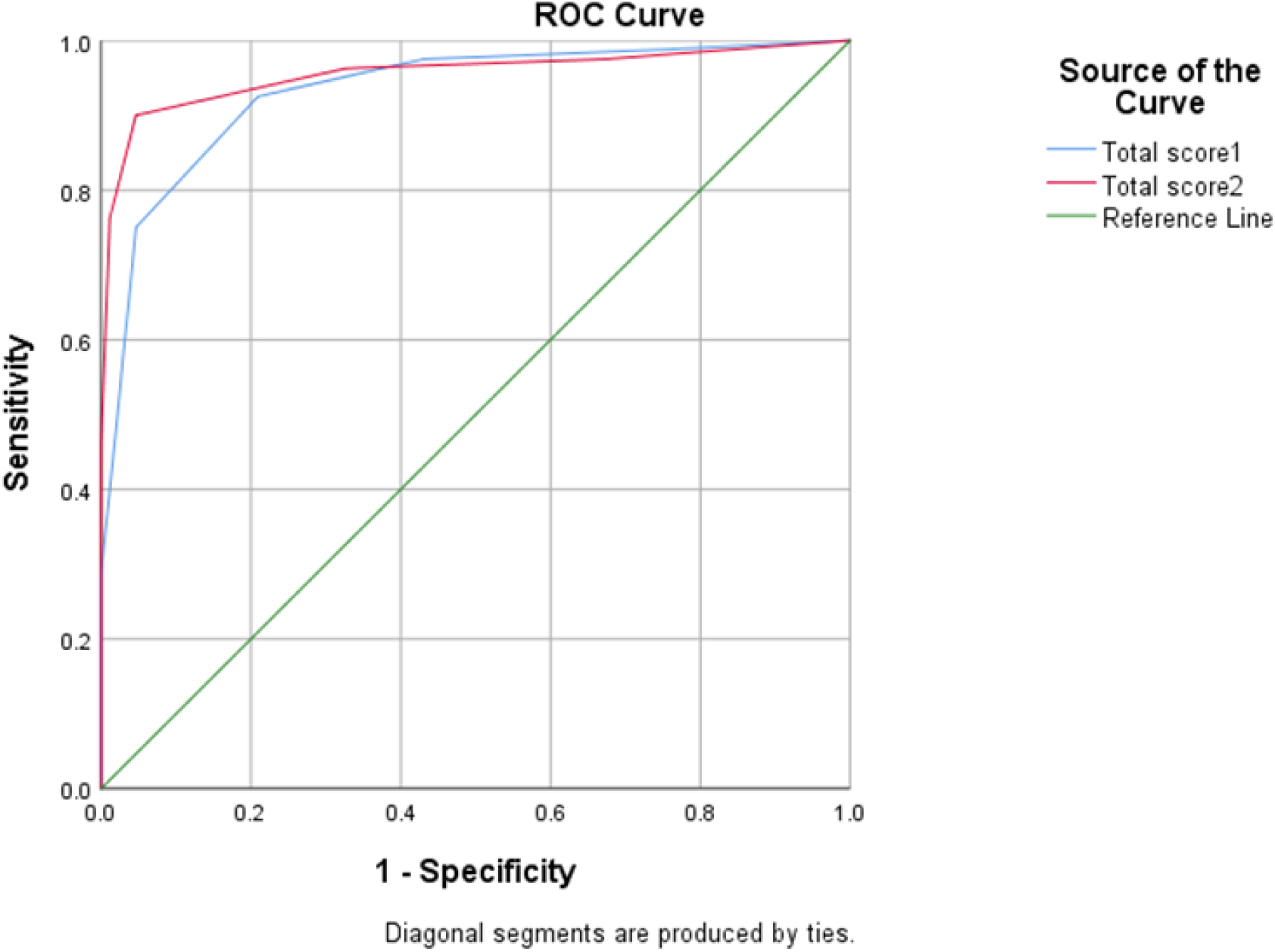
Receiving-operator characteristic (ROC) curve for the total score of DN4. AUC (Test 1) = 0.932 (95% CI: 0.894-0.971) AUC (Test 2) = 0.955 (95% CI: 0.921-0.990)

The sensitivity and specificity values for the two episodes of the test were found highest at the cut-off level of 4, which was also used in the original DN4 questionnaire. Similarly, other measures like Youden index, positive and negative predictive values, etc were also calculated (Table 6).

**Table 6.** Diagnostic Values of the DN4 questionnaire for the Discrimination of Neuropathic Pain.

| Test Result | Youden index | Sensitivity (%) | Specificity (%) | PPV (%) | NPV (%) | PLR (%) | NLR (%) |
| --- | --- | --- | --- | --- | --- | --- | --- |
| Test 1 | 0.7 | 75.0 | 95.3 | 93.8 | 80.4 | 15.96 | 0.26 |
| Test 2 | 0.75 | 76.2 | 98.8 | 98.4 | 81.7 | 63.5 | 0.24 |

PPV: Positive Predictive Value, NPV: Negative Predictive Value, PLR: Positive Likelihood Ratio, NLR: Negative Likelihood Ratio

## Discussion

This study was conducted at one of the academic institutes of Nepal among 166 patients who were referred to the pain clinic. The Nepali version of DN4 questionnaire was tested on the patients. The test-retest reliability was determined, along with AUC analysis and calculation of sensitivity, specificity, positive and negative predictive values with a cutoff value of 4, which was also originally used in French version (15). DN4 questionnaire was first created by French neuropathic team and was originally in French language. Due to its simplistic nature, the questionnaire has been translated into various other languages. However, translation can be a complicated process. The major hurdle is the fact that every language has its own unique nuances, grammatical rules, vocabulary, and so on. Rendering ideas from one language to another while also keeping the essence intact is a difficult task with a simple translation. The issues that could be encountered during translation of questionnaires were well discussed in the study by Seventer et al. (21). The authors described three situations: the original word can be literally translated, the word might need added explanation, or a different word that retains the intended meaning has to be used. For interpreting this questionnaire into Nepali language, we have conformed to the universal guidelines for translating questionnaires.

The first time we encountered a problem was while we were trying to restate questions 1 and 2 in Nepali. [Question 1: Does the pain have one or more of the following characteristics? Question 2: Is the pain associated with one or more of the following symptoms in the same area?] Due to the inherent discrepancies in the grammatical rules and lexicons of the two languages, achieving a direct rendering of the aforementioned questions proved infeasible. With the aid of two interpreters and clinical staff, we managed to come up with restatements that are close in meaning to the original questions, but also in line with the conventions of the Nepali language which Nepalese people can easily comprehend. Of all the items listed in the questionnaire, ‘cold pain’ was the most difficult term to properly translate. All other items except this term have Nepali equivalents. For this reason, we had to think of a phrase that conveyed the same meaning as ‘cold pain’. Most of the patients in the study actually believed that this kind of pain is aggravated by cold weather. Upon asking the question “Do you understand what it means?”, we identified this common misconception. For this reason, caution must be exercised while asking the patient about cold pain. In the Hindi version of DN4 questionnaire also, cold pain was present in only 14% of patients with neuropathic pain (7). In our study, only 17 % of patients with neuropathic pain had symptoms of cold pain. While interviewing twice using the Nepali version of the questionnaire, it showed a good test-retest reliability value (ICC=8). In the same way, the New Arabic version (22) of the questionnaire also showed excellent test-retest reliability (ICC>9). The test–retest intraclass correlation coefficient (95% CI) of the Japanese version was 0.827 (0.769–0.870) (9). Similarly, in our study, the AUC values for tests 1 and 2 were 0.932 with 95% CI (0.894-0.971) and 0.955 with 95% CI (0.921-0.990) respectively. The sensitivity and specificity of our version of the questionnaire were 75% and 95.3%, respectively, for test 1, and 76.2% and 98.8%, respectively, for test 2, which were the highest at the cut-off value 4 or more. This finding is in accordance with most of the validation studies (7,9,11). There are, however, validations done in Dutch (23) and New Arabic (22) languages, and both found the cutoff value at 5/10.

Based on our study, the most frequent complaints that patients with neuropathic pain had were burning pain, electrical sensation, tingling sensation, and numbness. When this is compared with other studies, we found variations (7,22,23). We believe that neuropathic pain consists of a collection of symptoms and signs that may also depend on the type of disease, so rather than focusing on any specific item for predicting neuropathic pain, one must consider the cutoff value. As per our study, a cutoff point of 4/10 is the best for predicting neuropathic pain.

Our sample consisted of both literate and illiterate patients (those who could communicate in Nepali language). However, we did not face any difficulties during the interviews, and the results were also very similar. This study can validate Nepali version of DN4 questionnaire so that it can be used as a standard tool to assess neuropathic pain among the Nepalese population. As the interview was conducted with the patients, this minimizes the reliability and validity issue that can occur in the case of a self-administered questionnaire. However, there were few limitations too. The patients were first seen by a pain physician who was not involved in the interview. He used clinical judgment for clinical diagnosis of pain. Other investigations could not be carried out due to financial constraints of the patients. Importantly, the institutional policy did not permit such investigations just for the sake of research. The same clinical diagnosis was used as reference for the calculation of the data. Besides, Nepal is a small country, but with multicultural and multilinguistic population. Thus, this questionnaire is only applicable to those who can communicate properly in Nepali language. There can be a problem of understanding among the participants as few words do not have an exact Nepali language with the same meaning, for instance, ‘cold pain’.

## Conclusion

The Nepalese version of DN4 questionnaire has been validated and can be used as a screening tool for predicting neuropathic pain in patients with chronic pain who can communicate well in Nepali.

## Supporting information

STARD checklist

Flow diagram

## Data Availability

The raw data can be made available upon genuine request.

## Acknowledgements

We would like to thank Mr. Laxman Rajbanshi (MA in Political Science) and Mr. Tirtha Ratna Shakya (MA) and Dr. Sarbottan Shrestha (Neurologist) for translating the questionnaire into Nepali and Mr. Sandesh Shakya (MA in English) for back translation. We would also like to thank the hospital and the patients who participated in the study

## Author Contributions

The corresponding author attests that all the following listed authors meet authorship criteria, and no others meeting the criteria have been omitted.

BMS: study concept and design, literature review, manuscript writing, review and editing

AS: literature review, manuscript review

AKP: data analysis, manuscript review

NS: data collection, literature review, manuscript review

BA: data collection, literature review, manuscript review

RG: data collection, literature review, manuscript review

SS: data analysis, manuscript writing and editing

## Patient consent for publication

Not required

## Ethical approval

The ethical approval for the study was obtained from the Institutional Review Committee of, Tribhuvan University, Institute of Medicine, Nepal (338(6-11)E^2^/075/76).

## Data availability statement

The raw data can be made available upon genuine request.

## Supplementary Material

The supplementary file consists of STARD checklist showing whether the required information has been included in manuscripts submitted for publication. It also includes the diagram showing the flow of participants through the study.

