## Supplementary material for "Nepalese version of Douleur Neuropathique 4 (DN4) questionnaire for Assessment of Neuropathic pain: A Validation Study": Flow diagram

Potentially eligible participants

n=216

Excluded due to various reasons (n=50)

Eligible participants

n=166

Index test (DN4 questionnaire)

n=166

Reference standard

Neuropathic pain (n=80)

Reference standard

Non-neuropathic pain (n=86)

Index test positive (Neuropathic pain)

n=64

Index test negative (Non-neuropathic pain)

n=102

**Supplemental Figure 1: Flow of the Participants through the Study**
